# Microbiological quality, antimicrobial resistance and hygiene-related risk factors of street-vended foods in Habiganj, Bangladesh: a spatially explicit cross-sectional study

**DOI:** 10.64898/2026.11.24.26368907

**Authors:** Sharmin Sultana Royal, Salauddin Yousup, Md. Abul Kalam Azad, Md. Shamim Uddin, Tasnim Zarin, FNU Nahiduzzaman, Md. Ali Asgar, Md. Ariful Islam, Md. Azizul Haque

## Abstract

Microbiological safety and antimicrobial resistance (AMR) in street-vended foods remain insufficiently characterized in Bangladesh’s secondary towns.

To assess bacterial contamination, antimicrobial susceptibility and hygiene-associated contamination factors in Habiganj municipality.

A cross-sectional study conducted during June - September surveyed 110 vendors at eight sites and examined 219 food, water and surface samples; 99 isolates underwent susceptibility testing against 13 antimicrobials. Analyses included multivariable logistic regression with vendor-clustered standard errors, Spearman correlation, principal component analysis (PCA) and hygiene-profile clustering.

Overall, 35.6% of samples contained *Escherichia coli*, *Salmonella* spp. or *Staphylococcus aureus*, and 41.6% were microbiologically unsatisfactory. Multivariable analysis identified higher contamination odds among vendors who never covered food (adjusted odds ratio [AOR] 5.16; p < 0.001), sometimes covered food (AOR 2.67; p = 0.035), or washed hands irregularly (AOR 2.08; p = 0.046), relative to consistent covering and regular handwashing, respectively. Hygiene-practice scores correlated inversely with mean total plate count (Spearman’s ρ = −0.26; p < 0.05), whereas food-holding time showed no correlation (ρ = 0.01; p = 0.897). The first two principal components explained 22.5% of hygiene-indicator variance; two weakly separated vendor clusters had comparable contamination prevalence (38.8% versus 34.2%; p = 0.616). Among isolates, 55.6% were multidrug resistant, resistance was highest to ampicillin (62.6%), and tetracycline–doxycycline exhibited the strongest co-resistance (φ = 0.73).

Frequent bacterial contamination and multidrug resistance warrant municipal interventions emphasizing consistent food covering, regular handwashing and integrated food-chain AMR surveillance.

## 1. Introduction

Foodborne disease remains a major and largely preventable cause of illness. The World Health Organization estimated that 31 foodborne hazards caused about 600 million illnesses and 420 000 deaths in 2010, with the heaviest per-capita burden in South-East Asia and Africa [1,2]. Diarrhoeal agents such as pathogenic *Escherichia coli* and non-typhoidal *Salmonella* account for a large share of this burden, while enterotoxigenic *Staphylococcus aureus* causes rapid-onset intoxication when contaminated food is held at warm temperatures [2,3]. Most of these illnesses are never investigated, so the hazards that cause them are rarely traced back to a specific food or place.

Street-vended foods are a large part of that unmeasured exposure. The Food and Agriculture Organization describes them as ready-to-eat foods and beverages prepared or sold by vendors in streets and other public places [4]. They provide inexpensive meals for students, labourers and commuters, and an income for vendors who often have little formal schooling or capital. The same features make them hazardous: vendors usually work without piped water, toilets or refrigeration, foods are held for hours at ambient temperature, and raw and cooked items share utensils and surfaces [5]. Reviews from Asia, Africa and Latin America have repeatedly recovered faecal indicators and pathogens from these foods, and contamination is generally linked to handling practices rather than to the raw ingredients alone [6].

Antimicrobial resistance (AMR) adds a second layer of risk. Bacterial AMR was associated with an estimated 4.95 million deaths in 2019, and South Asia was among the regions with the highest attributable mortality [7]. Antimicrobials are widely available without prescription in Bangladesh and are used extensively in poultry, aquaculture and livestock production. Resistant commensals and pathogens can therefore reach ready-to-eat foods through raw ingredients, water and the hands of food handlers. Street food is consumed without further cooking, so it gives resistant bacteria a direct route into the human gut, where resistance genes may be transferred to the resident microbiota. Food-chain surveillance is one of the pillars of the global action plan on AMR [7], but street foods are seldom included in national monitoring.

In Bangladesh, studies of street-food vendors have documented low levels of formal food-safety training, inadequate hand-washing facilities and frequent bare-hand handling of food and money [8,9]. Microbiological surveys have isolated *E. coli*, *S. aureus* and *Salmonella* spp. from fast foods and beverages, often with high resistance to penicillins and tetracyclines [4,5]. This evidence has three gaps. First, it is concentrated in Dhaka and a few divisional cities, while secondary towns, where municipal inspection capacity is thinner, are poorly represented. Second, most studies describe either vendor behaviour or laboratory findings, but rarely link the two at the level of the individual vendor, which is needed to identify modifiable risk factors. Third, AMR results are usually reported as drug-by-drug percentages, which hide how resistance accumulates across antimicrobial classes in the same isolate.

Habiganj is a district headquarters in Sylhet Division, close to the Dhaka–Sylhet highway. Its municipal centre has a dense street-food trade around the bus stand, markets, college and hospital, serving residents and a large transient population of travellers. We are not aware of any published study that has assessed the microbiological safety of street foods in this town. Food safety in Bangladesh is governed by the Food Safety Act 2013 and enforced by the Bangladesh Food Safety Authority, but routine inspection of mobile and roadside vendors is limited outside the largest cities. Mapping contamination at the level of individual vending sites could show inspectors where limited resources would have most effect, but such spatial analyses are rare for street food.

We therefore conducted a cross-sectional study of street-food vendors in Habiganj municipality with four objectives: (i) to estimate the microbial load and the prevalence of coliforms, *E. coli*, *Salmonella* spp. and *S. aureus* in ready-to-eat foods, water and food-contact surfaces; (ii) to describe the spatial distribution of contamination and multidrug resistance across vending sites; (iii) to identify vendor- and sample-level risk factors for pathogen contamination with a univariable screen followed by multivariable logistic regression; and (iv) to characterize the phenotypic AMR profiles of the recovered isolates, including co-resistance structure and multi-class resistance patterns.

## 2. Materials and methods

### 2.1 Study area and design

A cross-sectional study was conducted in Habiganj Paurashava (municipality), Habiganj Sadar Upazila, Sylhet Division, Bangladesh, between 15 June and 31 September 2026 (Fig 1). Eight vending sites with high footfall were selected: Bus Stand, Chowdhury Bazar, College Road, Court Station, Hospital Road, Kalibari Road, New Market and Shaistaganj Road. All vendors who were selling ready-to-eat food at these sites on the survey days and who gave consent were enrolled. In total, 110 vendors took part, and one to six samples (median two) were collected from each vendor, giving 219 samples.

**Fig 1.**
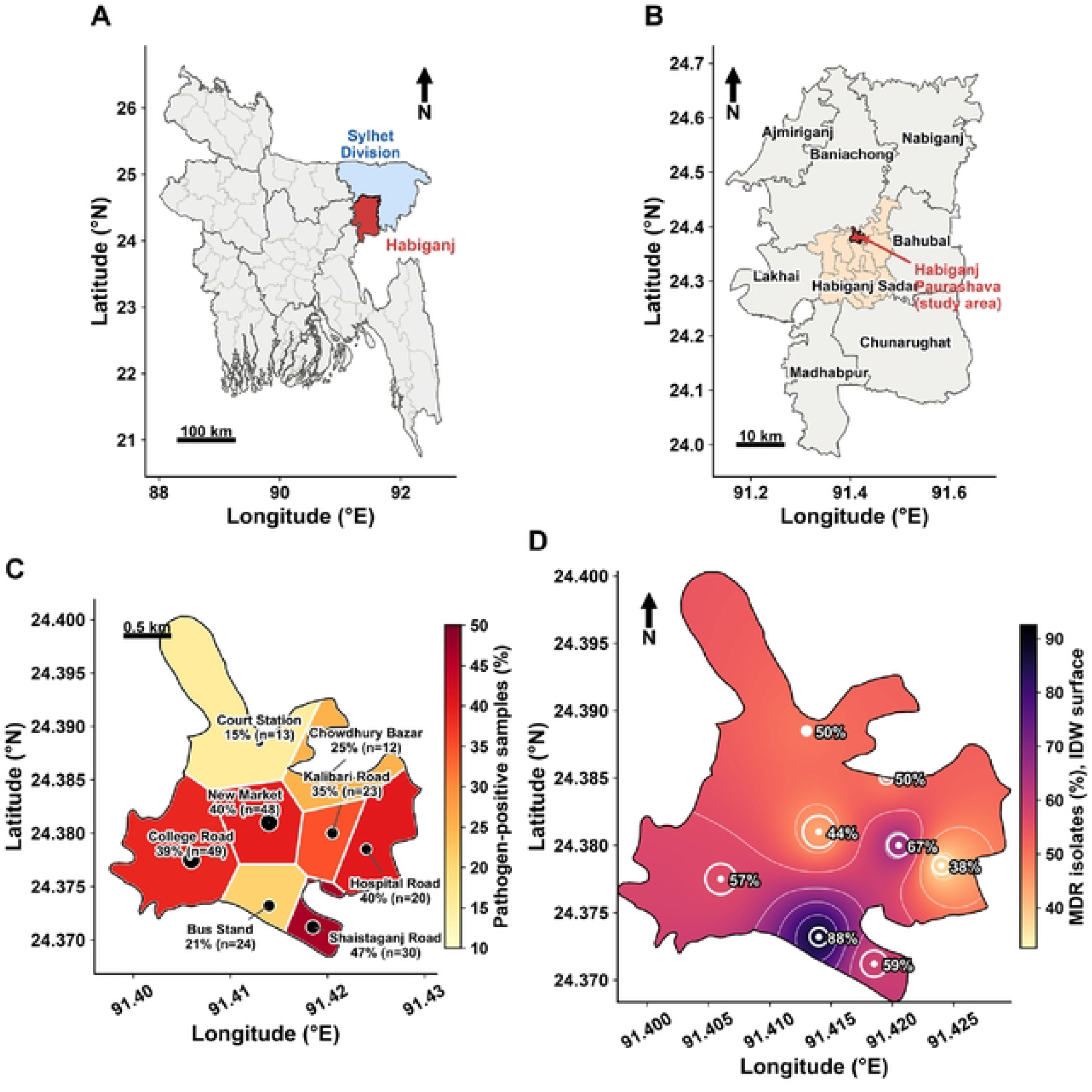
Study area and spatial distribution of contamination and multidrug resistance. (A) Bangladesh with Sylhet Division (blue) and Habiganj District (red). (B) Upazilas of Habiganj District, showing Habiganj Sadar Upazila (shaded) and Habiganj Paurashava, the study area (red). (C) Thiessen polygons of the eight vending sites clipped to the municipal boundary and shaded by the percentage of samples positive for at least one target pathogen; point size is proportional to the number of samples. (D) Inverse-distance-weighted surface of the percentage of MDR isolates; rings are proportional to the number of isolates, and labels give site-level MDR percentages. Boundaries: GADM v4.l.

### 2.2 Vendor questionnaire and hygiene observation

Each vendor completed a face-to-face structured interview in Bangla covering socio-demographic characteristics, selling experience, food-safety training, trade licence and inspection history. Hygiene conditions were recorded by direct observation with a checklist covering water source, glove use, hand washing, utensil cleaning, handling of money and food, food covering, waste disposal, soap and hand-washing station availability, covering and daily replacement of stored water, refrigeration, separation of raw and cooked utensils, stall type, drainage, fly exposure and distance to the nearest waste point. Knowledge (0–15), attitude (0–10) and practice (0–15) were scored and summed into an overall KAP score (0–40). Knowledge and practice were classified as poor (≤ 5), moderate (6–10) or good (≥ 11), and attitude as negative (≤ 3), neutral (4–7) or positive (≥ 8). Each vendor was also given a composite hygiene-risk rating (low, medium or high) based on the observed conditions.

### 2.3 Sample collection

Ready-to-eat food samples (chotpoti, burger, grilled chicken, fruit juice, samosa and fuchka) were purchased as served to customers. Water used for juice preparation or for washing utensils was collected in sterile bottles, and swabs were taken from vendors’ hands, utensils, cutting boards, knives and serving plates with sterile cotton swabs moistened in nutrient broth. At collection, the time of day, ambient temperature, food-holding time and condition (ambient, hot-held or chilled), use of previous-day leftovers, rainfall in the previous 24 h and addition of water after cooking were recorded. Samples were placed in sterile bags or containers, transported to the laboratory in an insulated box with ice packs and processed on the day of collection.

### 2.4 Microbiological analysis

Ten grams of solid food or 10 mL of liquid sample was homogenised in 90 mL of nutrient broth (NB), and swabs were vortexed in 10 mL of NB. Ten-fold serial dilutions were prepared, and 0.1 mL of appropriate dilutions was spread in duplicate on plate count agar (PCA). Plates were incubated at 37 °C for 24-48 h, plates bearing 30-300 colonies were counted, and the total plate count (TPC) was expressed as log_10_ CFU/g or mL.

For selective isolation, aliquots of each homogenate were enriched in three broths at 37 °C for 18-24 h: NB for coliforms and *E. coli*, trypticase soy broth (TSB) for *S. aureus*, and selenite broth (SB) for *Salmonella* spp. The NB enrichment was streaked onto eosin methylene blue (EMB) agar; lactose-fermenting colonies were recorded as coliforms, and colonies with a green metallic sheen were taken as presumptive *E. coli*. The TSB enrichment was streaked onto mannitol salt agar (MSA), where yellow colonies surrounded by yellow zones were taken as presumptive *S. aureus*. The SB enrichment was streaked onto *Salmonella*–*Shigella* (SS) agar, where colourless colonies with black centres were taken as presumptive *Salmonella* spp. All plates were incubated at 37 °C for 24 h. Presumptive colonies were purified by sub-culture and confirmed by Gram staining and conventional biochemical tests. Samples were classified as satisfactory, marginal or unsatisfactory by combining TPC with the detection of indicator organisms and pathogens, following the Health Protection Agency guidelines for ready-to-eat foods [10]. One isolate of each organism per positive sample was kept for susceptibility testing.

### 2.5 Antimicrobial susceptibility testing

Susceptibility of the 99 isolates (36 *E. coli*, 20 *Salmonella* spp. and 43 *S. aureus*) was tested by the Kirby-Bauer disc diffusion method on Mueller-Hinton agar (MHA). Bacterial suspensions were adjusted to 0.5 McFarland turbidity and swabbed evenly over the agar surface, antimicrobial discs were applied, and plates were incubated at 37 °C for 16-18 h. Inhibition-zone diameters were interpreted as susceptible (S), intermediate (I) or resistant (R) according to Clinical and Laboratory Standards Institute guidance [14]. Thirteen antimicrobials from five classes were tested: penicillins (ampicillin, amoxicillin, penicillin G), aminoglycosides (gentamicin, streptomycin, amikacin), fluoroquinolones (ciprofloxacin, enrofloxacin, levofloxacin), tetracyclines (tetracycline, doxycycline) and phenicols (chloramphenicol, florfenicol). Intermediate results were counted as non-resistant. Multidrug resistance (MDR) was defined as resistance to at least one agent in three or more classes [11]. The multiple antibiotic resistance (MAR) index was calculated as the number of antimicrobials to which an isolate was resistant divided by the number tested (13); values above 0.2 were taken to indicate origin from a high-risk source with heavy antimicrobial use [12].

### 2.6 Statistical analysis

Categorical variables were summarised as counts and percentages with Wilson score 95% confidence intervals (CIs) [13], and continuous variables as mean ± standard deviation or median (interquartile range). Proportions were compared with Pearson’s chi-square test and distributions with the Kruskal–Wallis test. Correlations between vendor-level variables (KAP scores, age, experience, distance to waste, customers per day, mean TPC and proportion of pathogen-positive samples) were assessed with Spearman’s rank correlation.

The outcome for risk-factor analysis was sample-level positivity for at least one target pathogen (*E. coli*, *Salmonella* spp. or *S. aureus*). Thirty-seven candidate variables describing the sample, the vending site and the vendor were examined. Continuous variables were categorised for interpretability (vendor age, selling experience, food-holding time, ambient temperature, distance to waste and daily customers). Because only four vendors (11 samples, none positive) had a negative attitude, attitude was collapsed into positive versus neutral/negative to avoid complete separation. Each variable was first examined in univariable logistic regression, and its overall significance was assessed with the likelihood-ratio test. Variables with p < 0.20 were entered together into a multivariable logistic regression model [14]. Because several samples came from the same vendor, standard errors were estimated with a cluster-robust (sandwich) estimator with vendors as clusters. As a sensitivity analysis, the same model was fitted with generalised estimating equations (GEE) using an exchangeable working correlation [15]. Multicollinearity was checked with variance inflation factors (VIF), calibration with the Hosmer–Lemeshow test and discrimination with the area under the receiver operating characteristic curve (AUC). Risk factors were identified from the adjusted odds ratios (AORs), their 95% CIs and p-values in the multivariable model, with p < 0.05 considered significant.

For AMR, the percentage of resistant isolates by species and antimicrobial was displayed as a heat map with rows ordered by average-linkage hierarchical clustering. Pairwise co-resistance between antimicrobials was measured with the phi (φ) coefficient. Combinations of resistance classes within isolates were shown with an UpSet plot [21], which displays the size of each exact class combination and is easier to read than a Venn diagram when more than three sets are involved.

For spatial analysis, administrative boundaries were taken from the GADM database version 4.1 [16], and the approximate centre of each sampling site was georeferenced within the Habiganj Paurashava boundary. Site-level pathogen prevalence was mapped onto Thiessen (Voronoi) polygons clipped to the municipal boundary, and the percentage of MDR isolates was interpolated across the municipality by inverse distance weighting (power 2) [17].

To describe vendor hygiene profiles without reference to the outcome, 15 hygiene indicators (seven ordinal practice items scored 0–2 and eight binary infrastructure, training and inspection items) were standardised and summarised by principal component analysis (PCA) [18]. Vendors were grouped by k-means clustering (50 random starts), with the number of clusters (2–6) chosen by the mean silhouette width [19]. The relationship between food-holding time and TPC was explored with LOWESS smoothing and Spearman correlation.

Analyses were performed in Python 3.14 with pandas 3.0, statsmodels 0.14.6, SciPy 1.18, scikit-learn 1.9, GeoPandas 1.1.4 and Matplotlib 3.11. Two-sided p < 0.05 was considered statistically significant.

### 2.7 Ethical considerations

The study was conducted in accordance with the ethical protocols of Habiganj Agricultural University. All applicable ethical procedures were followed throughout the study. Informed consent was obtained from each vendor before the interview and sample purchase. Vendor identities were coded and are not reported.

## 3. Results

### 3.1 Characteristics of vendors

Of the 110 vendors, 89 (80.9%) were male; their mean age was 41.4 ± 12.0 years and median selling experience was 7 years (IQR 3–10). Most had primary (32.7%) or secondary (39.1%) schooling. Only 31 vendors (28.2%) had received food-safety training, 22 (20.0%) held a trade licence and 25 (22.7%) had been inspected in the preceding 12 months. Hygiene infrastructure was limited: soap was available at 35.5% of stalls, a hand-washing station at 42.7% and refrigeration at 19.1%. Food was always covered by 24.5% of vendors and never by 29.1%, and 55.5% handled money and food with the same hands. The full socio-demographic, hygiene and KAP profile of the vendors is given in Table 1.

**Table 1.**
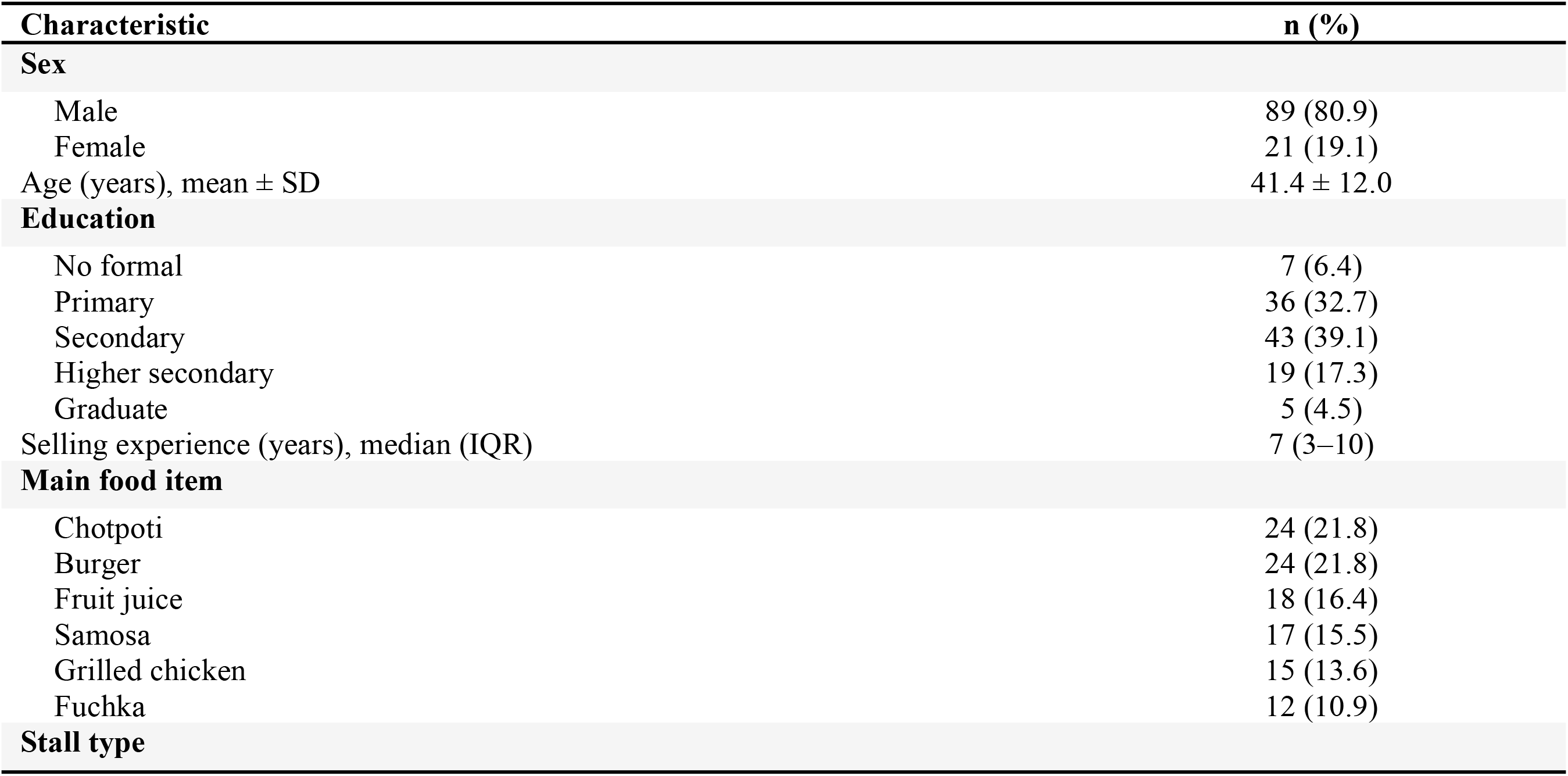
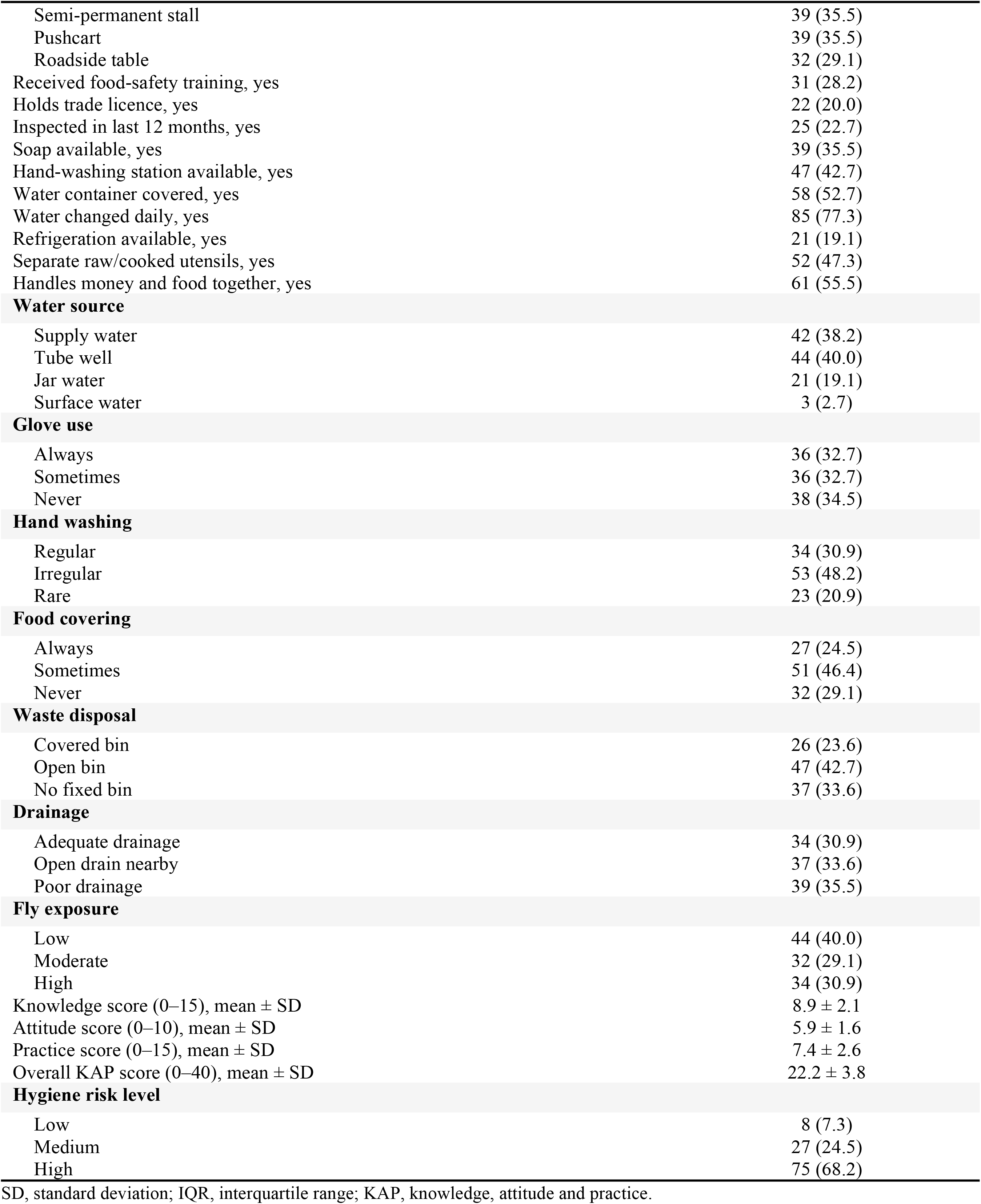
Socio-demographic, hygiene and KAP characteristics of street-food vendors in Habiganj municipality (n = 110).

| Characteristic | n (%) |
|---|---|
| **Sex** |  |
| Male | 89 (80.9) |
| Female | 21 (19.1) |
| Age (years), mean ± SD | 41.4 ± 12.0 |
| **Education** |  |
| No formal | 7 (6.4) |
| Primary | 36 (32.7) |
| Secondary | 43 (39.1) |
| Higher secondary | 19 (17.3) |
| Graduate | 5 (4.5) |
| Selling experience (years), median (IQR) | 7 (3–10) |
| **Main food item** |  |
| Chotpoti | 24 (21.8) |
| Burger | 24 (21.8) |
| Fruit juice | 18 (16.4) |
| Samosa | 17 (15.5) |
| Grilled chicken | 15 (13.6) |
| Fuchka | 12 (10.9) |
| **Stall type** |  |
| Semi-permanent stall | 39 (35.5) |
| Pushcart | 39 (35.5) |
| Roadside table | 32 (29.1) |
| Received food-safety training, yes | 31 (28.2) |
| Holds trade licence, yes | 22 (20.0) |
| Inspected in last 12 months, yes | 25 (22.7) |
| Soap available, yes | 39 (35.5) |
| Hand-washing station available, yes | 47 (42.7) |
| Water container covered, yes | 58 (52.7) |
| Water changed daily, yes | 85 (77.3) |
| Refrigeration available, yes | 21 (19.1) |
| Separate raw/cooked utensils, yes | 52 (47.3) |
| Handles money and food together, yes | 61 (55.5) |
| **Water source** |  |
| Supply water | 42 (38.2) |
| Tube well | 44 (40.0) |
| Jar water | 21 (19.1) |
| Surface water | 3 (2.7) |
| **Glove use** |  |
| Always | 36 (32.7) |
| Sometimes | 36 (32.7) |
| Never | 38 (34.5) |
| **Hand washing** |  |
| Regular | 34 (30.9) |
| Irregular | 53 (48.2) |
| Rare | 23 (20.9) |
| **Food covering** |  |
| Always | 27 (24.5) |
| Sometimes | 51 (46.4) |
| Never | 32 (29.1) |
| **Waste disposal** |  |
| Covered bin | 26 (23.6) |
| Open bin | 47 (42.7) |
| No fixed bin | 37 (33.6) |
| **Drainage** |  |
| Adequate drainage | 34 (30.9) |
| Open drain nearby | 37 (33.6) |
| Poor drainage | 39 (35.5) |
| **Fly exposure** |  |
| Low | 44 (40.0) |
| Moderate | 32 (29.1) |
| High | 34 (30.9) |
| Knowledge score (0–15), mean ± SD | 8.9 ± 2.1 |
| Attitude score (0–10), mean ± SD | 5.9 ± 1.6 |
| Practice score (0–15), mean ± SD | 7.4 ± 2.6 |
| Overall KAP score (0–40), mean ± SD | 22.2 ± 3.8 |
| **Hygiene risk level** |  |
| Low | 8 (7.3) |
| Medium | 27 (24.5) |
| High | 75 (68.2) |

SD, standard deviation; IQR, interquartile range; KAP, knowledge, attitude and practice.

### 3.2 Microbiological quality of food, water and surfaces

Mean TPC across all 219 samples was 4.49 ± 1.03 log_10_ CFU/g or mL (range 2.10–6.74), and 17 samples (7.8%) reached at least 6 log_10_ CFU (Fig 2A). Median TPC was highest in grilled chicken (5.09 log_10_), cutting-board swabs (4.96) and fruit juice (4.77), but differences between sample types were not significant (Kruskal–Wallis p = 0.328). Overall, 41.6% of samples were unsatisfactory, 34.7% marginal and 23.7% satisfactory; the unsatisfactory share ranged from 14% in utensil swabs to 54% in fruit juice (Fig 2B) and differed between sample categories (food 46.0%, water 40.0%, swabs 19.4%; p = 0.022).

**Fig 2.**
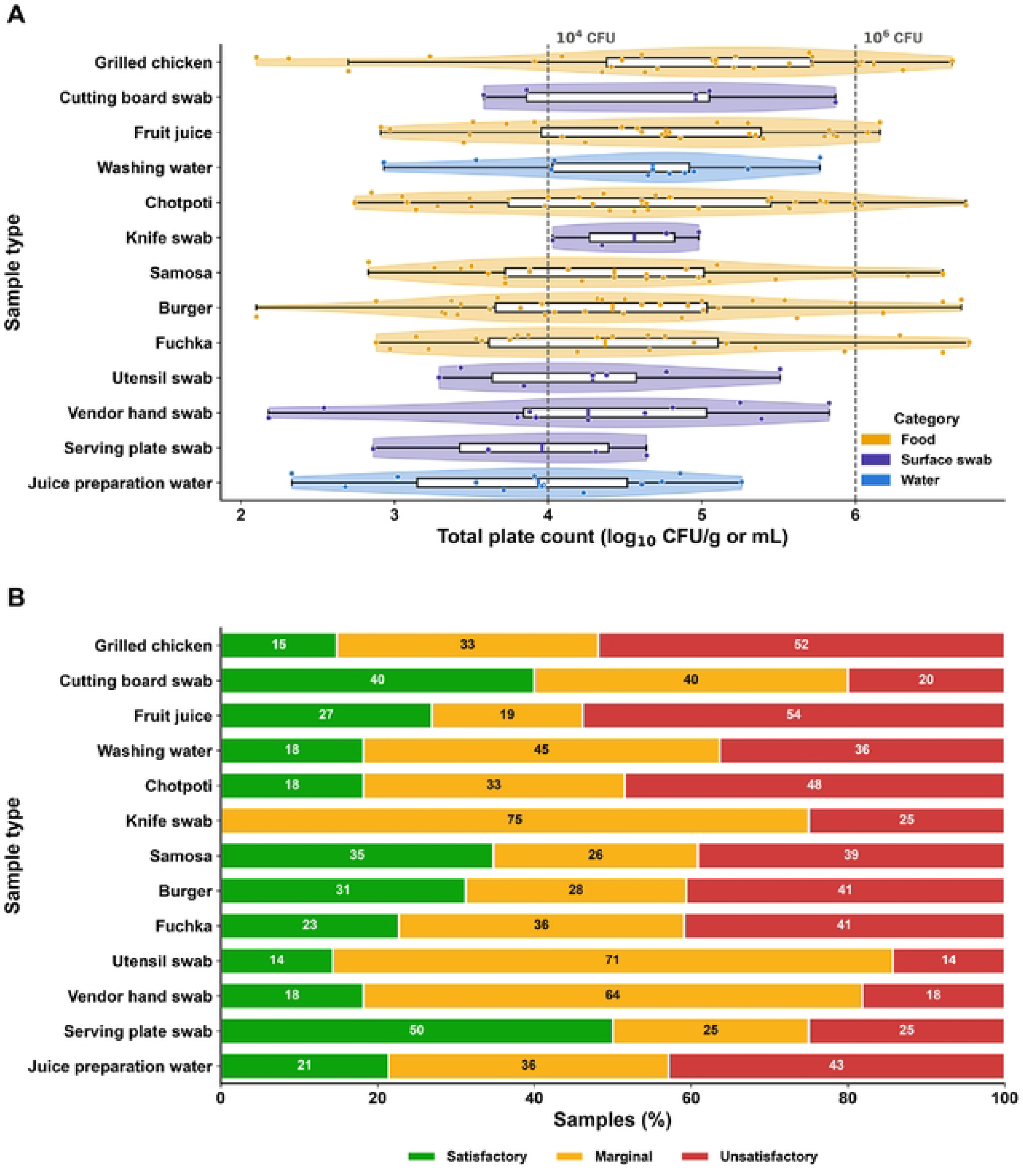
Microbial load and acceptability of street-vended food, water and surface samples. (A) Distribution of total plate count (log_10_ CFU/g or mL) by sample type, shown as violin, box (median and IQR) and individual points, coloured by sample category; dashed lines mark I0^4^ and I0^6^ CFU. (B) Percentage of satisfactory, marginal and unsatisfactory samples by sample type, in the same order as panel A.

Coliforms were detected in 33.3% of samples, *E. coli* in 16.4%, *S. aureus* in 16.4% and *Salmonella* spp. in 5.9%. At least one target pathogen was present in 78 samples (35.6%; 95% CI 29.6–42.2): 35.0% of foods, 52.0% of water samples and 25.8% of surface swabs (p = 0.119). Juice-preparation water had the highest positivity (57.1%), driven by *E. coli* (42.9%), followed by knife swabs (50.0%), washing water (45.5%) and chotpoti (42.4%) (Fig 3A). Among foods, *S. aureus* was most frequent in burgers (21.9%) and *Salmonella* spp. in chotpoti (9.1%). Coliforms were recovered from 45.5% of vendor hand swabs, although only one hand swab yielded a target pathogen. The TPC and the prevalence of each organism by sample category and type are summarised in Table 2.

**Fig 3.**
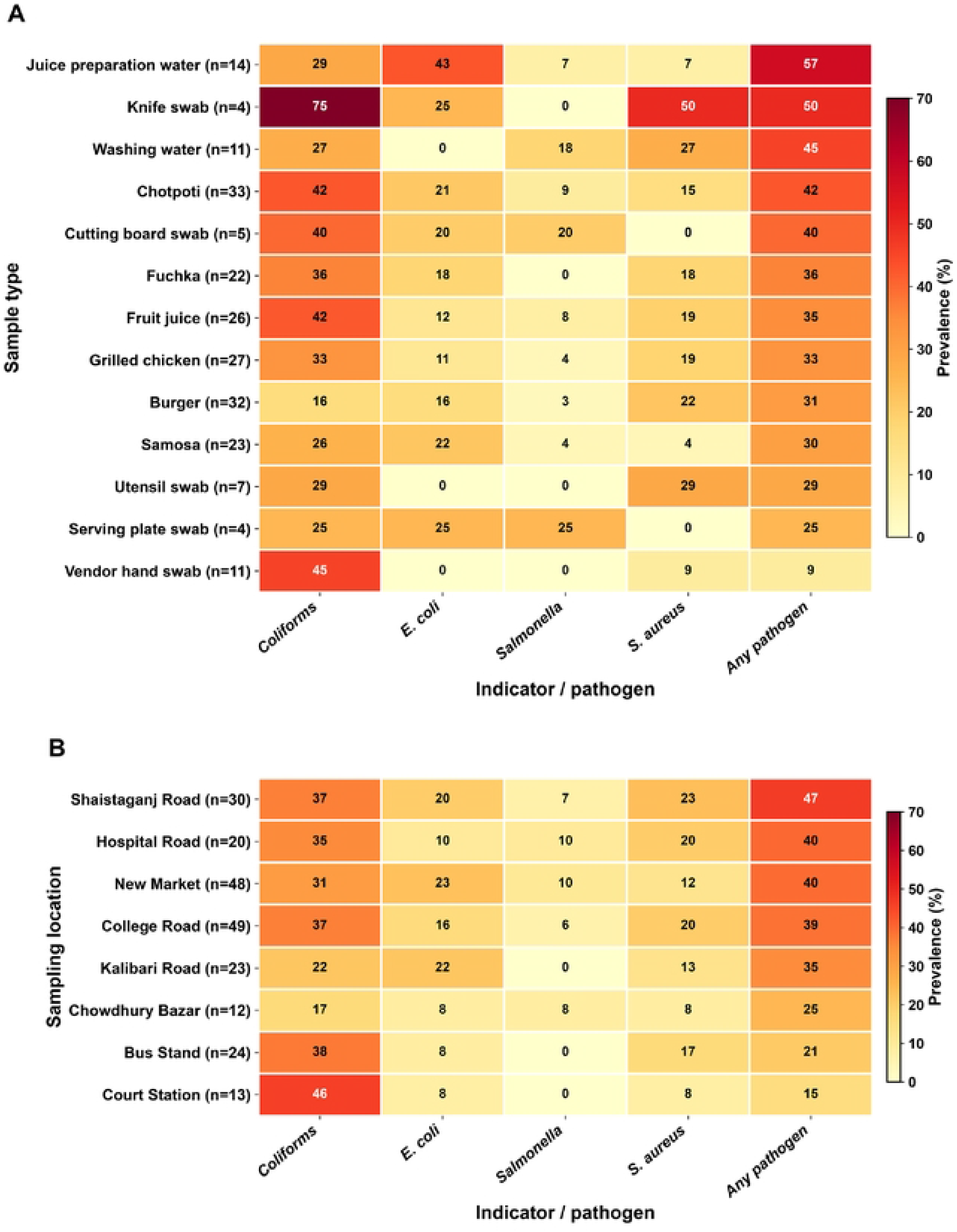
Prevalence of coliforms and target pathogens. Heat maps of the percentage of positive samples (A) by sample type and (B) by sampling location, ordered by positivity for any target pathogen *(E.coli, Salmonella* spp. or S. *aureus).* Numbers in cells are percentages.

**Table 2.**
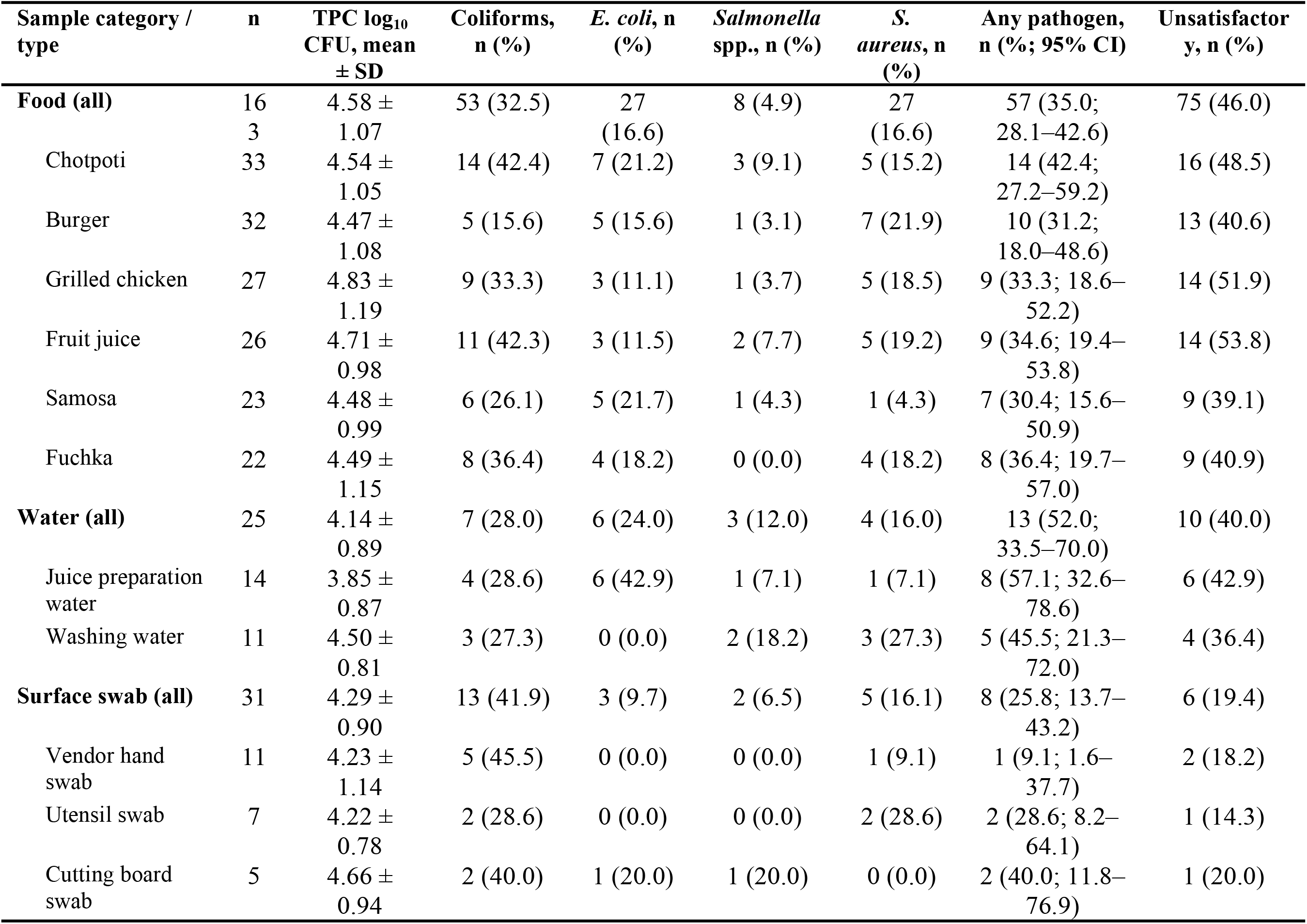
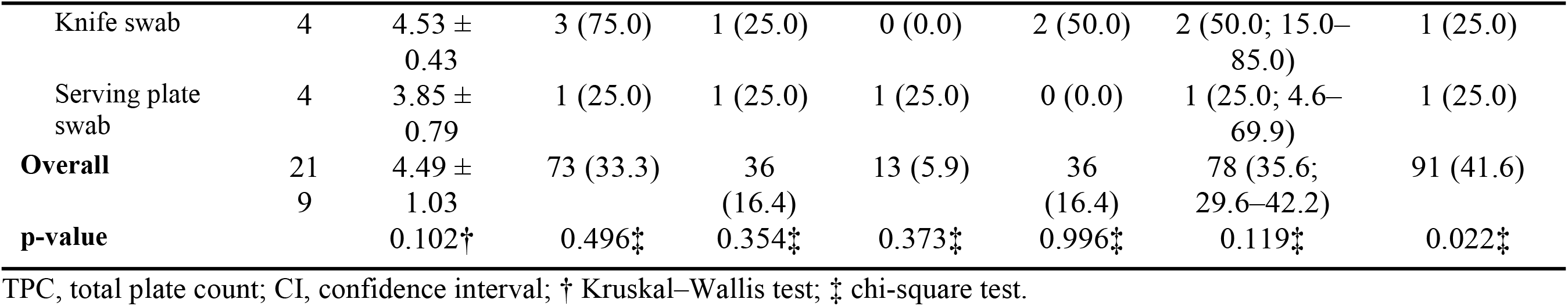
Total plate count and prevalence of coliforms and target pathogens by sample category and type (n = 219).

### 3.3 Spatial distribution

Pathogen positivity varied from 15.4% at Court Station to 46.7% at Shaistaganj Road, with New Market (39.6%), Hospital Road (40.0%) and College Road (38.8%) also above the overall mean (Fig 1C, Fig 3B); the difference between sites was not statistically significant (p = 0.378). Mean TPC, however, differed between sites (Kruskal–Wallis p = 0.004) and was highest at New Market (4.91 log_10_). The interpolated MDR surface showed a hotspot in the south of the municipality around the Bus Stand, where 7 of 8 isolates (87.5%) were MDR, and lower values on Hospital Road (3/8, 37.5%) (Fig 1D). These site-level MDR proportions rest on small numbers of isolates and did not differ significantly (p = 0.486).

### 3.4 Knowledge, attitude and practice

The mean knowledge, attitude and practice scores were 8.9 ± 2.1 of 15, 5.9 ± 1.6 of 10 and 7.4 ± 2.6 of 15, respectively, and the overall KAP score was 22.2 ± 3.8 of 40 (Fig 4A). Seventy per cent of vendors had moderate knowledge and 25.5% good knowledge, but 80.9% held a neutral attitude and 25.5% had poor practice; only 14.5% were rated as having good practice (Fig 4B). Two-thirds of vendors (68.2%) were classified as high hygiene risk. Practice scores correlated weakly and negatively with the vendor’s mean TPC (ρ = −0.26, p < 0.05), as did vendor age (ρ = −0.31), whereas knowledge and attitude were not correlated with practice or with contamination (Fig 4C).

**Fig 4.**
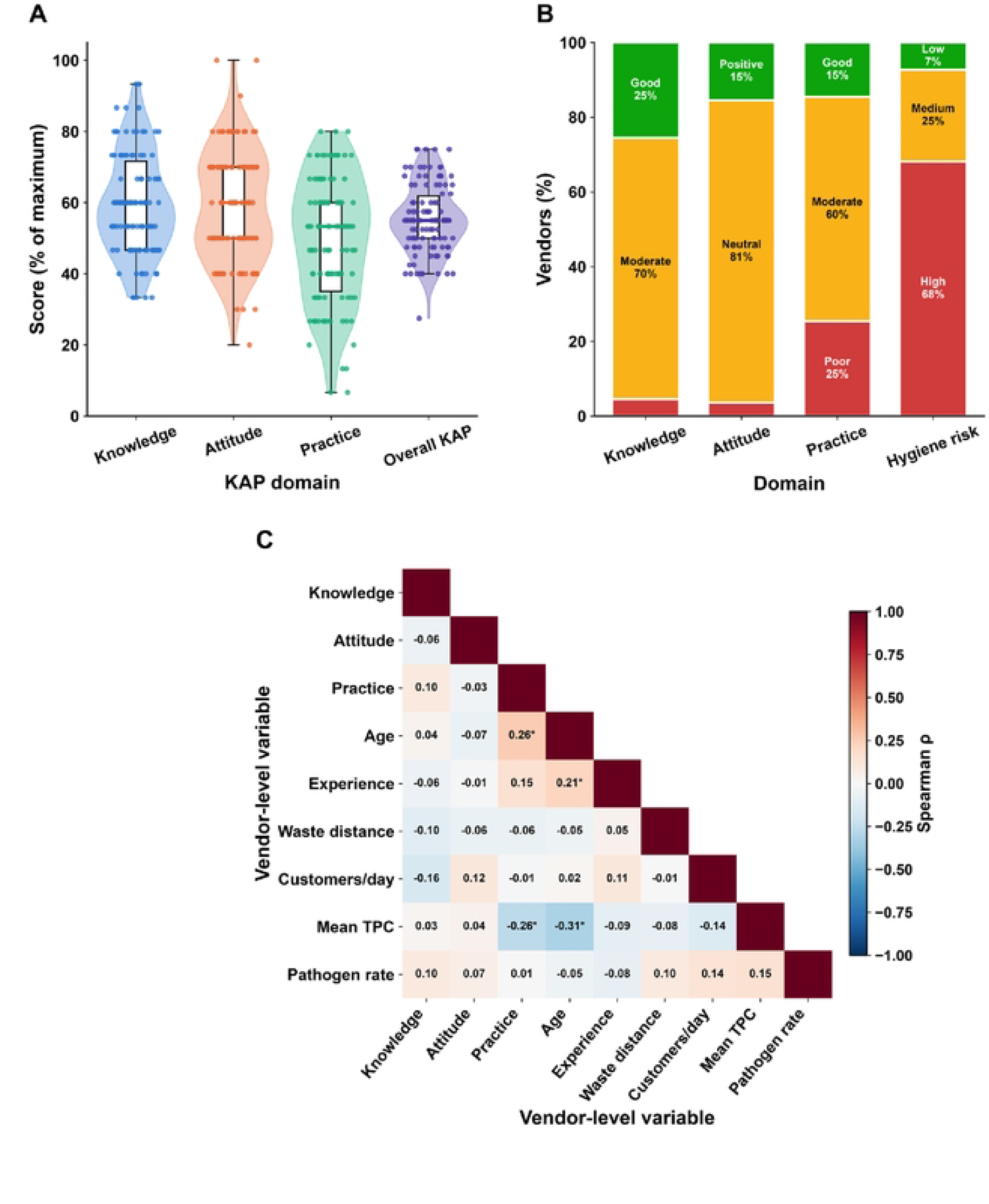
Food-safety knowledge, attitude and practice of street-food vendors (n = I IO). (A) Domain scores expressed as a percentage of the maximum score. (B) Distribution of vendors across knowledge, attitude, practice and hygiene-risk categories. (C) Spearman correlation matrix of vendor-level variables, including each vendor’s mean TPC and proportion of pathogen-positive samples; *p < 0.05.

### 3.5 Univariable analysis of factors associated with contamination

Of the 37 candidate variables, nine passed the univariable screen at p < 0.20: sample category, food-safety training, water source, glove use, hand washing, utensil cleaning, food covering, attitude level and hand-washing station availability. In crude analysis, never covering food (COR 2.77, 95% CI 1.28–6.02), rare hand washing (COR 2.21, 1.03–4.75) and the absence of a hand-washing station (COR 1.83, 1.05–3.21) were associated with higher odds of contamination, whereas tube-well or surface water (COR 0.49, 0.26–0.91) and occasional glove use (COR 0.37, 0.18–0.77) were associated with lower odds. Attitude level lost significance after it was collapsed to two levels (p = 0.81) and was not carried forward. The remaining variables were not associated with contamination (all p ≥ 0.20): sampling location, time of collection, ambient temperature, holding time and condition, previous-day leftovers, recent rain, water added after cooking, vendor age, sex, education and experience, trade licence, handling of money with food, waste disposal, knowledge and practice levels, soap availability, covering and daily replacement of stored water, refrigeration, separate utensils, stall type, drainage, fly exposure, distance to waste, inspection and number of customers. The crude odds ratios of the eight variables retained for multivariable analysis are presented in Table 3.

**Table 3.**
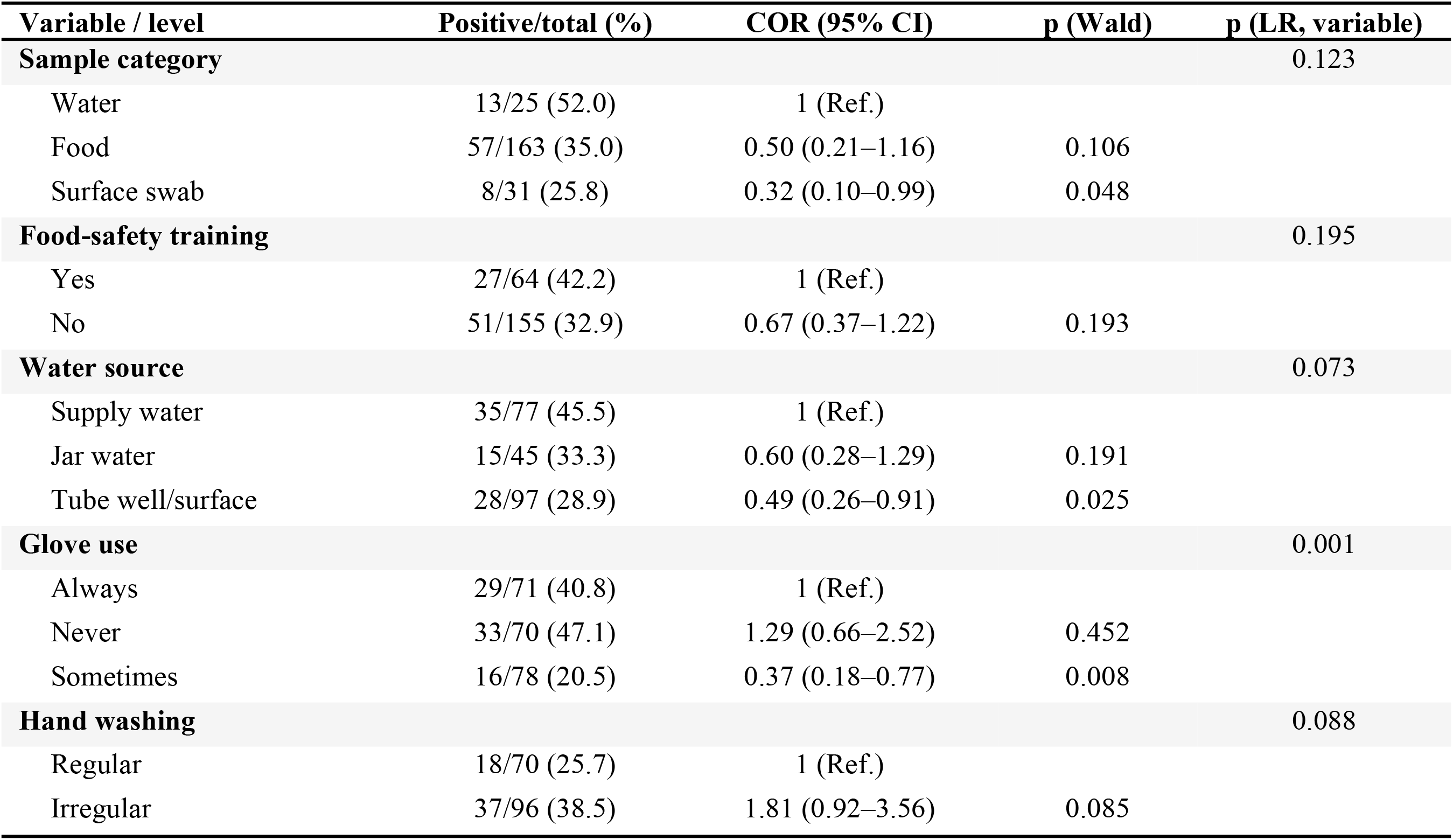
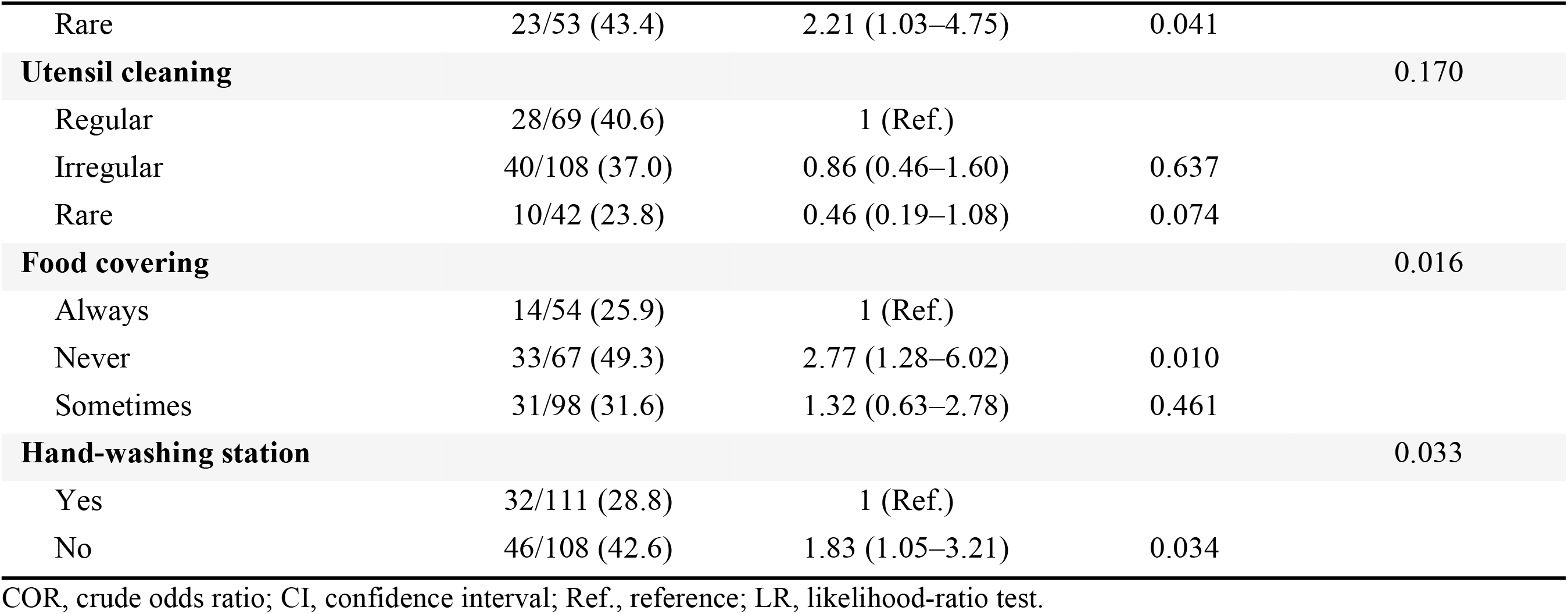
Univariable logistic regression of factors associated with pathogen positivity: variables retained for multivariable analysis (p < 0.20).

| **Variable / level** | **Positive/total (%)** | **COR (95% CI)** | **p (Wald)** | **p (LR, variable)** |
|---|---|---|---|---|
| **Sample category** |  |  |  | 0.123 |
| Water | 13/25 (52.0) | 1 (Ref.) |  |  |
| Food | 57/163 (35.0) | 0.50 (0.21–1.16) | 0.106 |  |
| Surface swab | 8/31 (25.8) | 0.32 (0.10–0.99) | 0.048 |  |
| **Food-safety training** |  |  |  | 0.195 |
| Yes | 27/64 (42.2) | 1 (Ref.) |  |  |
| No | 51/155 (32.9) | 0.67 (0.37–1.22) | 0.193 |  |
| **Water source** |  |  |  | 0.073 |
| Supply water | 35/77 (45.5) | 1 (Ref.) |  |  |
| Jar water | 15/45 (33.3) | 0.60 (0.28–1.29) | 0.191 |  |
| Tube well/surface | 28/97 (28.9) | 0.49 (0.26–0.91) | 0.025 |  |
| **Glove use** |  |  |  | 0.001 |
| Always | 29/71 (40.8) | 1 (Ref.) |  |  |
| Never | 33/70 (47.1) | 1.29 (0.66–2.52) | 0.452 |  |
| Sometimes | 16/78 (20.5) | 0.37 (0.18–0.77) | 0.008 |  |
| **Hand washing** |  |  |  | 0.088 |
| Regular | 18/70 (25.7) | 1 (Ref.) |  |  |
| Irregular | 37/96 (38.5) | 1.81 (0.92–3.56) | 0.085 |  |
| Rare | 23/53 (43.4) | 2.21 (1.03–4.75) | 0.041 |  |
| **Utensil cleaning** |  |  |  | 0.170 |
| Regular | 28/69 (40.6) | 1 (Ref.) |  |  |
| Irregular | 40/108 (37.0) | 0.86 (0.46–1.60) | 0.637 |  |
| Rare | 10/42 (23.8) | 0.46 (0.19–1.08) | 0.074 |  |
| **Food covering** |  |  |  | 0.016 |
| Always | 14/54 (25.9) | 1 (Ref.) |  |  |
| Never | 33/67 (49.3) | 2.77 (1.28–6.02) | 0.010 |  |
| Sometimes | 31/98 (31.6) | 1.32 (0.63–2.78) | 0.461 |  |
| **Hand-washing station** |  |  |  | 0.033 |
| Yes | 32/111 (28.8) | 1 (Ref.) |  |  |
| No | 46/108 (42.6) | 1.83 (1.05–3.21) | 0.034 |  |

COR, crude odds ratio; CI, confidence interval; Ref., reference; LR, likelihood-ratio test.

### 3.6 Multivariable risk factors for contamination

In the multivariable model (219 samples, 78 events), food covering and hand washing were independent risk factors. Samples from vendors who never covered their food had five times higher odds of pathogen contamination than those from vendors who always covered it (AOR 5.16, 95% CI 1.98–13.40, p < 0.001), and covering only sometimes also raised the odds (AOR 2.67, 1.07–6.67, p = 0.035). Irregular hand washing roughly doubled the odds compared with regular hand washing (AOR 2.08, 1.01–4.26, p = 0.046); the estimate for rare hand washing was in the same direction but imprecise (AOR 1.34, 0.54–3.31). Three variables showed inverse associations: tube-well or surface water compared with municipal supply water (AOR 0.40, 0.20–0.81, p = 0.010), occasional compared with constant glove use (AOR 0.28, 0.10–0.78, p = 0.014) and no training compared with training (AOR 0.45, 0.21–0.95, p = 0.036). Water samples tended to be more often positive than food or swab samples, but not significantly after adjustment. The model was significant overall (likelihood-ratio p < 0.001), well calibrated (Hosmer–Lemeshow p = 0.84) and moderately discriminating (AUC 0.74; McFadden pseudo-R² 0.15), and showed no collinearity (maximum VIF 2.25). GEE estimates were almost identical in size and direction. The adjusted odds ratios from both models are shown in Table 4.

**Table 4.** Multivariable logistic regression of risk factors for pathogen positivity (n = 219 samples from 110 vendors; 78 positive).

| Variable / level | AOR (95% CI) | p | GEE AOR (95% CI) | p (GEE) |
|---|---|---|---|---|
| **Sample category** |  |  |  |  |
| Water | 1 (Ref.) |  | 1 (Ref.) |  |
| Food | 0.40 (0.14–1.13) | 0.084 | 0.39 (0.15–1.04) | 0.059 |
| Surface swab | 0.26 (0.06–1.08) | 0.065 | 0.25 (0.06–0.99) | 0.048 |
| **Food-safety training** |  |  |  |  |
| Yes | 1 (Ref.) |  | 1 (Ref.) |  |
| No | 0.45 (0.21–0.95) | 0.036 | 0.45 (0.22–0.92) | 0.029 |
| **Water source** |  |  |  |  |
| Supply water | 1 (Ref.) |  | 1 (Ref.) |  |
| Jar water | 0.69 (0.29–1.63) | 0.392 | 0.70 (0.31–1.59) | 0.397 |
| Tube well/surface | 0.40 (0.20–0.81) | 0.010 | 0.40 (0.20–0.77) | 0.006 |
| **Glove use** |  |  |  |  |
| Always | 1 (Ref.) |  | 1 (Ref.) |  |
| Never | 1.84 (0.83–4.08) | 0.131 | 1.97 (0.92–4.21) | 0.081 |
| Sometimes | 0.28 (0.10–0.78) | 0.014 | 0.30 (0.11–0.81) | 0.018 |
| **Hand washing** |  |  |  |  |
| Regular | 1 (Ref.) |  | 1 (Ref.) |  |
| Irregular | 2.08 (1.01–4.26) | 0.046 | 2.24 (1.13–4.44) | 0.021 |
| Rare | 1.34 (0.54–3.31) | 0.526 | 1.35 (0.56–3.23) | 0.500 |
| **Utensil cleaning** |  |  |  |  |
| Regular | 1 (Ref.) |  | 1 (Ref.) |  |
| Irregular | 0.88 (0.43–1.81) | 0.726 | 0.83 (0.41–1.67) | 0.605 |
| Rare | 1.97 (0.62–6.27) | 0.252 | 1.80 (0.58–5.54) | 0.307 |
| **Food covering** |  |  |  |  |
| Always | 1 (Ref.) |  | 1 (Ref.) |  |
| Never | 5.16 (1.98–13.40) | <0.001 | 5.23 (2.10–13.04) | <0.001 |
| Sometimes | 2.67 (1.07–6.67) | 0.035 | 2.78 (1.15–6.73) | 0.023 |
| **Hand-washing station** |  |  |  |  |
| Yes | 1 (Ref.) |  | 1 (Ref.) |  |
| No | 1.44 (0.77–2.72) | 0.257 | 1.46 (0.79–2.70) | 0.224 |

AOR, adjusted odds ratio; CI, confidence interval; Ref., reference; GEE, generalised estimating equations.

### 3.7 Antimicrobial resistance

Resistance was most common to ampicillin (62.6%), tetracycline (55.6%), amoxicillin (54.5%) and penicillin G (51.5%), and least common to gentamicin (13.1%), amikacin (17.2%) and chloramphenicol (20.2%) (Fig 5). Ampicillin resistance was lower in *Salmonella* spp. (30.0%) than in *E. coli* (69.4%) or *S. aureus* (72.1%) (p = 0.003), and ciprofloxacin resistance was highest in *S. aureus* (37.2%) and lowest in *Salmonella* (5.0%) (p = 0.025). Florfenicol resistance was more frequent in *E. coli* (38.9%) than in the other species (p = 0.059).

**Fig 5.**
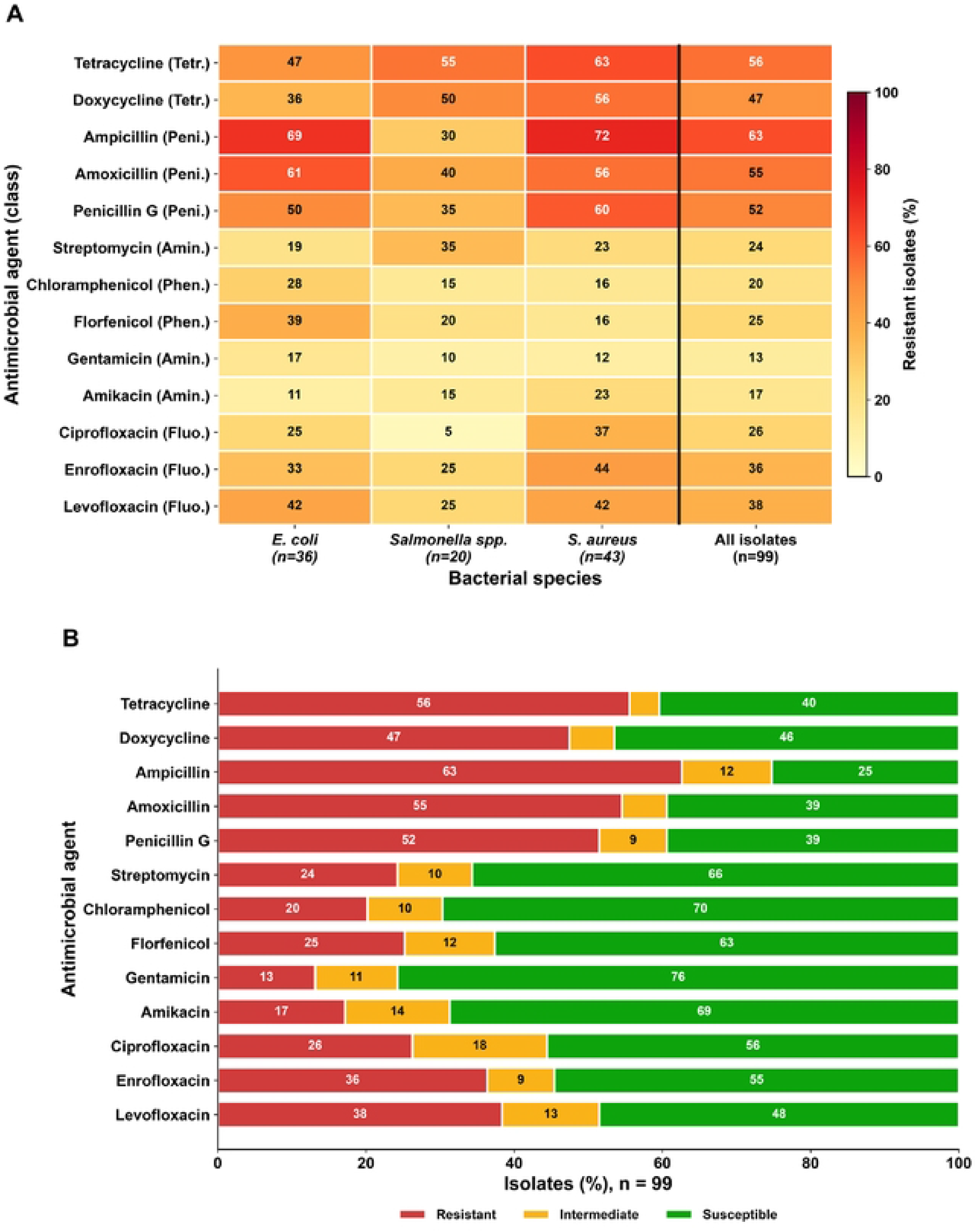
Antimicrobial resistance of *E. coli, Salmonella* spp. and S. *aureus* isolates (n = 99). (A) Percentage of resistant isolates by species and antimicrobial; rows are ordered by hierarchical clustering of species-level resistance and labelled with the antim.icrobial class (Peni., penicillins; Amin., aminoglycosides; Fluo., fluoroquinolones; Tetr., tetracyclines; Phen., phenicols). (B) Percentage of all isolates classified as resistant, intermediate or susceptible.

Only three isolates (3.0%) were susceptible to all five classes. Fifty-five isolates (55.6%; 95% CI 45.7–65.0) were MDR, including 60.5% of *S. aureus*, 58.3% of *E. coli* and 40.0% of *Salmonella* spp. (p = 0.288), and four isolates were resistant to all five classes. The UpSet plot (Fig 6) showed 25 distinct class combinations. The most frequent was penicillin–fluoroquinolone–tetracycline resistance (14 isolates, 10 of them *S. aureus*), followed by penicillin–tetracycline–phenicol resistance (9 isolates, mostly *E. coli*). Penicillin resistance was present in 80 isolates and was part of every one of the five most common combinations.

**Fig 6.**
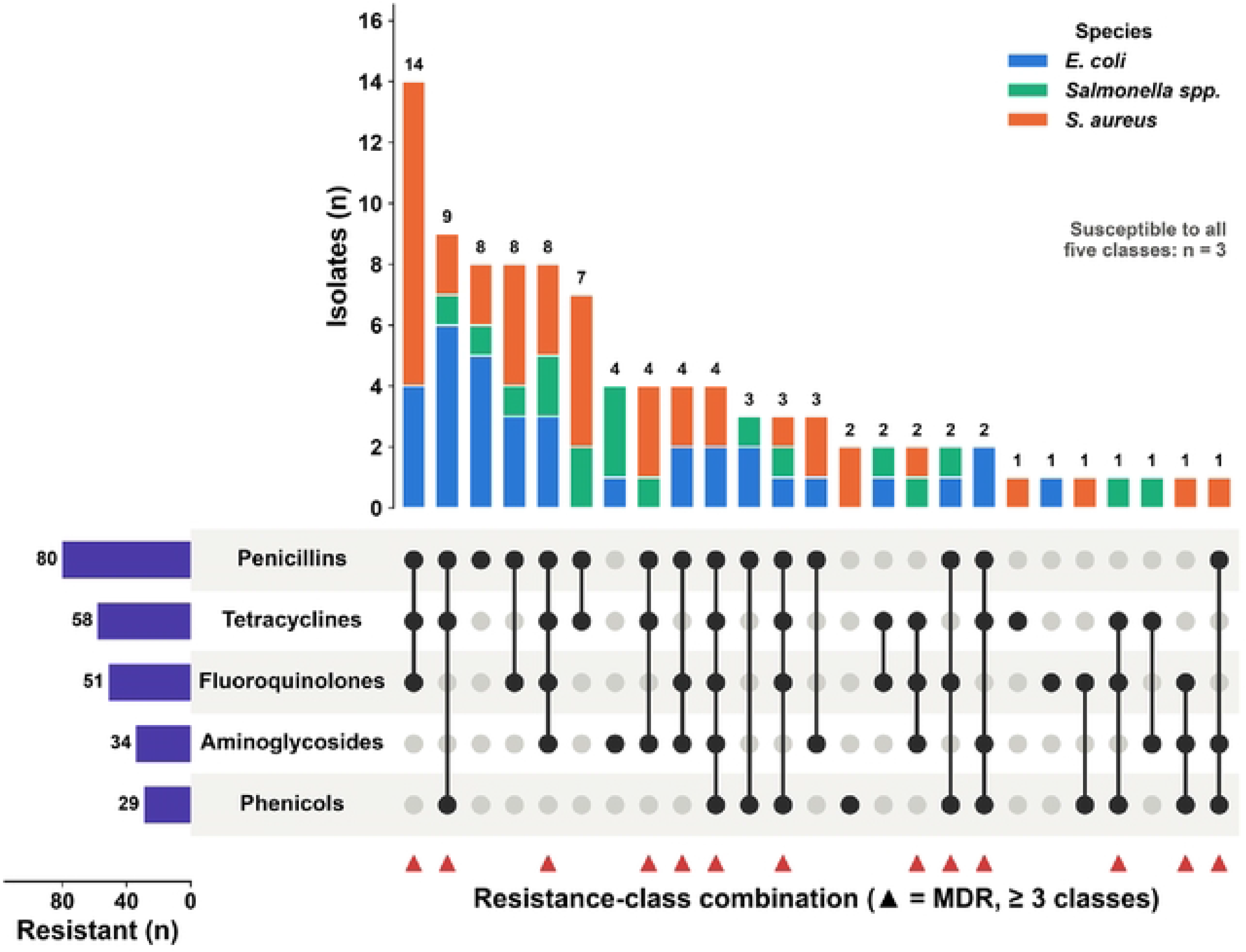
UpSet plot of antimicrobial-class resistance combinations among 96 isolates resistant to at least one class. The left bars give the number of isolates resistant to each class. Each column of the matrix is one exact combination of classes (filled dots), and the bar above it gives the number of isolates with that combination, stacked by species. Red triangles mark combinations that meet the MDR definition (three or more classes).

The mean MAR index was 0.36 ± 0.17 and 80.8% of isolates exceeded 0.2 (Fig 7A). MAR differed between species (Kruskal–Wallis p = 0.031), being highest in *S. aureus* (0.40 ± 0.15) and lowest in *Salmonella* spp. (0.28 ± 0.18). MDR prevalence was similar in isolates from food (54.2%), water (53.3%) and surfaces (66.7%) (Fig 7B; p = 0.710). Co-resistance was strongest between drugs of the same class: tetracycline–doxycycline (φ = 0.73), ampicillin–penicillin G (φ = 0.71), ciprofloxacin–levofloxacin (φ = 0.66), chloramphenicol–florfenicol (φ = 0.63) and ciprofloxacin–enrofloxacin (φ = 0.55), whereas cross-class correlations were weak (|φ| < 0.25) (Fig 7C). The resistance of each species to every antimicrobial, together with MDR prevalence and MAR index, is given in Table 5.

**Fig 7.**
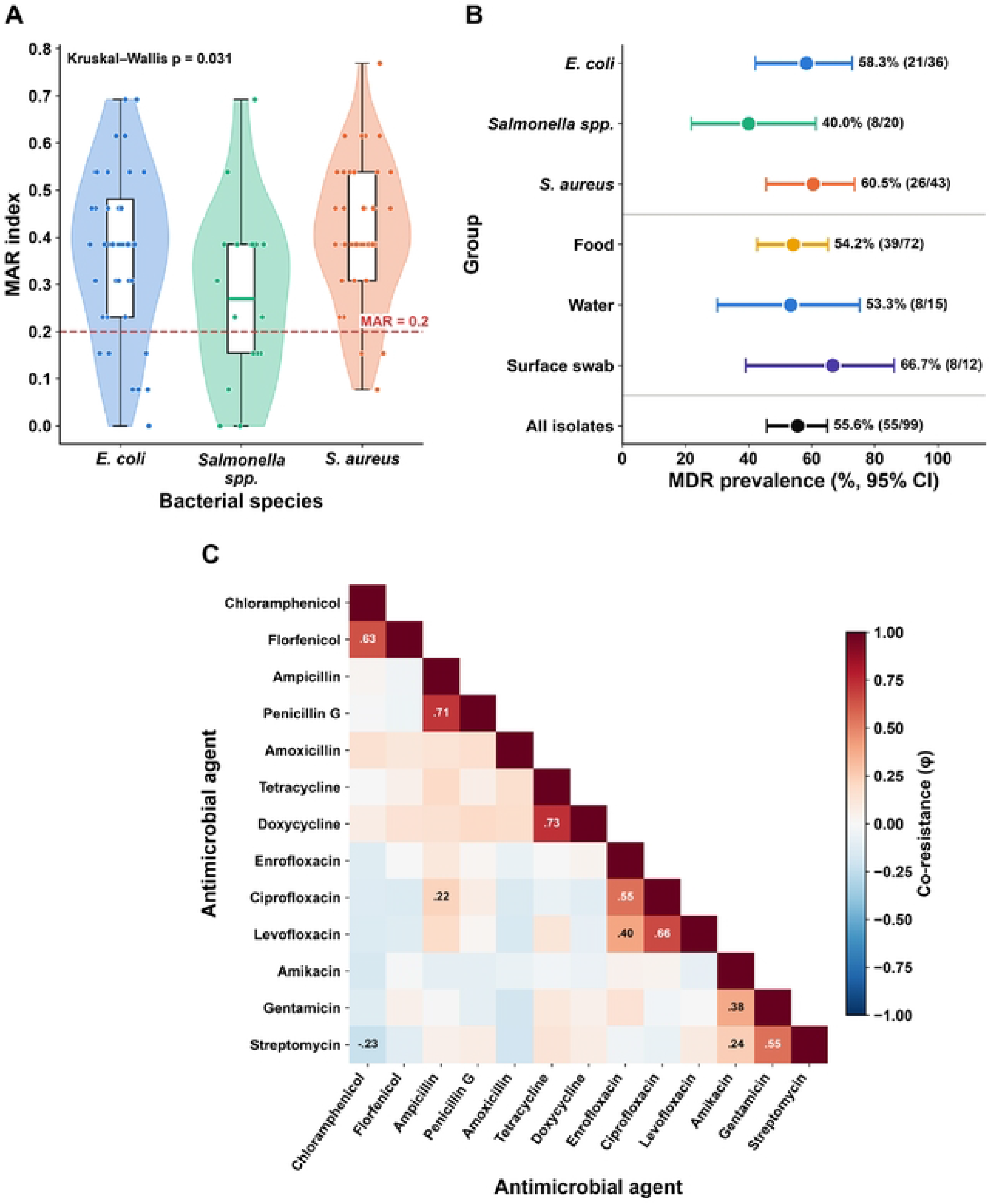
Multiple antibiotic resistance, multidrug resistance and co-resistance. (A) MAR index by species; the dashed line marks the 0.2 high-risk threshold. (B) MOR prevalence with Wilson 95% confidence intervals by species, sample source and overall. (C) Phi (cp) co-resistance coefficients between antimicrobials, ordered by hierarchical clustering; values with lφI ≥ 0.20 are printed.

**Table 5.** Antimicrobial resistance, multidrug resistance and MAR index of *E. coli*, *Salmonella* spp. and *S. aureus* isolates (n = 99).

| **Antimicrobial** | **Class** | ***E. coli* (n=36), R n (%; 95% CI)** | ***Salmonella* spp. (n=20), R n (%; 95% CI)** | ***S. aureus* (n=43), R n (%; 95% CI)** | **All (n=99), R n (%; 95% CI)** | **p** |
|---|---|---|---|---|---|---|
| Ampicillin | Penicillins | 25 (69.4; 53.1–82.0) | 6 (30.0; 14.5–51.9) | 31 (72.1; 57.3–83.3) | 62 (62.6; 52.8–71.5) | 0.003 |
| Amoxicillin | Penicillins | 22 (61.1; 44.9–75.2) | 8 (40.0; 21.9–61.3) | 24 (55.8; 41.1–69.6) | 54 (54.5; 44.8–64.0) | 0.307 |
| Penicillin G | Penicillins | 18 (50.0; 34.5–65.5) | 7 (35.0; 18.1–56.7) | 26 (60.5; 45.6–73.6) | 51 (51.5; 41.8–61.1) | 0.166 |
| Gentamicin | Aminoglycosides | 6 (16.7; 7.9–31.9) | 2 (10.0; 2.8–30.1) | 5 (11.6; 5.1–24.5) | 13 (13.1; 7.8–21.2) | 0.722 |
| Streptomycin | Aminoglycosides | 7 (19.4; 9.8–35.0) | 7 (35.0; 18.1–56.7) | 10 (23.3; 13.2–37.7) | 24 (24.2; 16.9–33.5) | 0.420 |
| Amikacin | Aminoglycosides | 4 (11.1; 4.4–25.3) | 3 (15.0; 5.2–36.0) | 10 (23.3; 13.2–37.7) | 17 (17.2; 11.0–25.8) | 0.347 |
| Ciprofloxacin | Fluoroquinolones | 9 (25.0; 13.8–41.1) | 1 (5.0; 0.9–23.6) | 16 (37.2; 24.4–52.1) | 26 (26.3; 18.6–35.7) | 0.025 |
| Enrofloxacin | Fluoroquinolones | 12 (33.3; 20.2–49.7) | 5 (25.0; 11.2–46.9) | 19 (44.2; 30.4–58.9) | 36 (36.4; 27.6–46.2) | 0.302 |
| Levofloxacin | Fluoroquinolones | 15 (41.7; 27.1–57.8) | 5 (25.0; 11.2–46.9) | 18 (41.9; 28.4–56.7) | 38 (38.4; 29.4–48.2) | 0.387 |
| Tetracycline | Tetracyclines | 17 (47.2; 32.0–63.0) | 11 (55.0; 34.2–74.2) | 27 (62.8; 47.9–75.6) | 55 (55.6; 45.7–65.0) | 0.382 |
| Doxycycline | Tetracyclines | 13 (36.1; 22.5–52.4) | 10 (50.0; 29.9–70.1) | 24 (55.8; 41.1–69.6) | 47 (47.5; 37.9–57.2) | 0.211 |
| Chloramphenicol | Phenicols | 10 (27.8; 15.8–44.0) | 3 (15.0; 5.2–36.0) | 7 (16.3; 8.1–30.0) | 20 (20.2; 13.5–29.2) | 0.363 |
| Florfenicol | Phenicols | 14 (38.9; 24.8–55.1) | 4 (20.0; 8.1–41.6) | 7 (16.3; 8.1–30.0) | 25 (25.3; 17.7–34.6) | 0.059 |
| MDR (≥3 classes) |  | 21 (58.3; 42.2–72.9) | 8 (40.0; 21.9–61.3) | 26 (60.5; 45.6–73.6) | 55 (55.6; 45.7–65.0) | 0.288 |
| MAR index, mean ± SD |  | 0.37 ± 0.18 | 0.28 ± 0.18 | 0.40 ± 0.15 | 0.36 ± 0.17 | 0.031 |
| MAR index > 0.2 |  | 29 (80.6) | 13 (65.0) | 38 (88.4) | 80 (80.8) | 0.090 |

CI, confidence interval; MDR, multidrug resistant (≥ 3 classes); MAR, multiple antibiotic resistance.

### 3.8 Vendor hygiene profiles

The first two principal components explained 12.7% and 9.8% of the variance in the 15 hygiene indicators, showing that hygiene behaviours did not collapse onto a single dimension. The silhouette criterion favoured two clusters, but separation was weak (mean silhouette 0.11) (Fig 8A). Cluster 1 (31 vendors) consisted mainly of trained (90%) and recently inspected (65%) vendors with better soap and hand-washing-station availability; cluster 2 (79 vendors) was almost entirely untrained (4%) and uninspected (6%). Pathogen positivity (38.8% vs 34.2%; p = 0.616) and the unsatisfactory proportion (41.8% vs 41.4%) were nearly the same in the two clusters (Fig 8B). Food-holding time was not correlated with TPC (ρ = 0.01, p = 0.897), and the LOWESS curve for hot-held foods lay below that for foods held at ambient temperature (Fig 8C).

**Fig 8.**
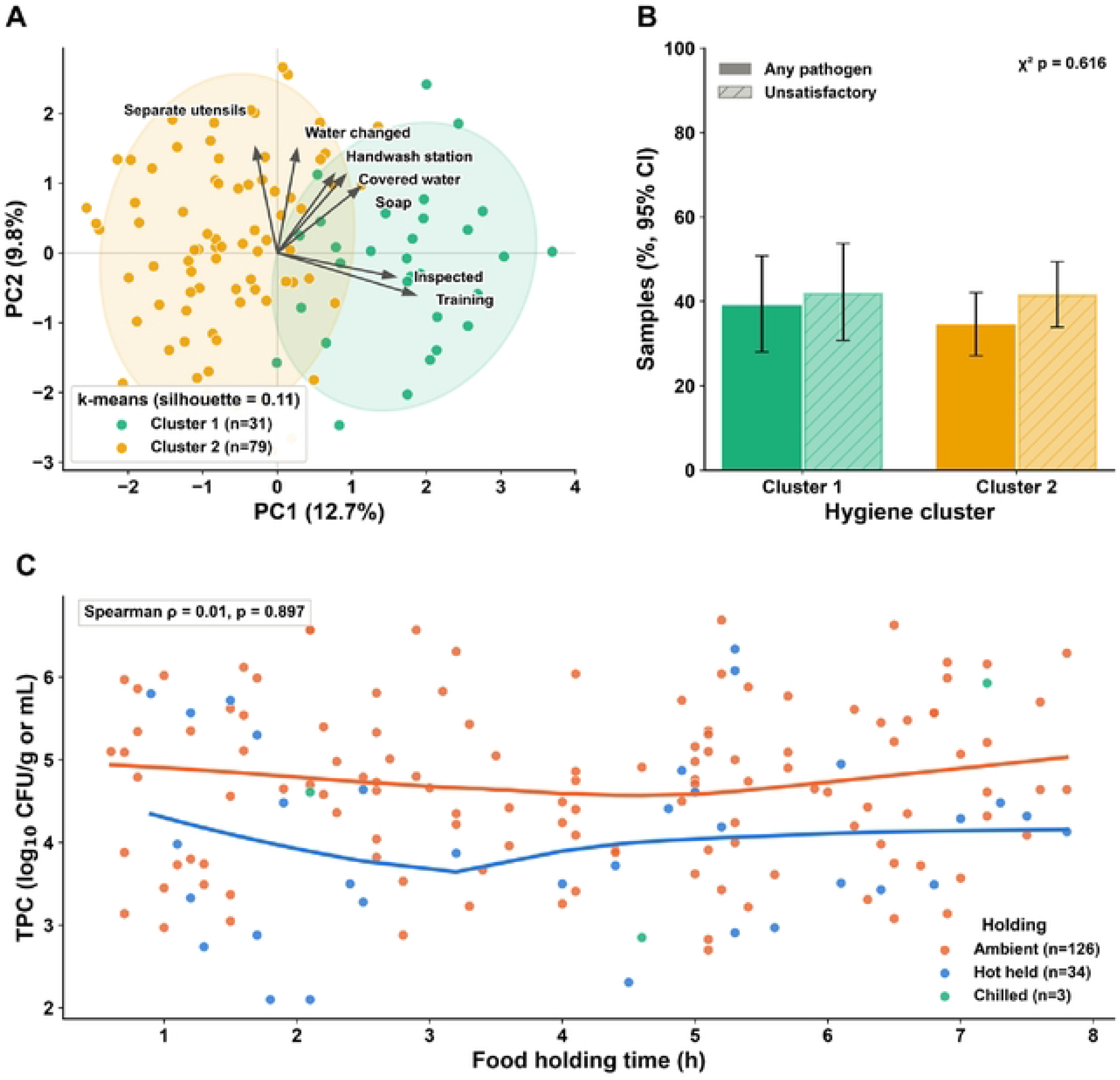
Vendor hygiene profiles and food holding. (A) Principal component analysis of I5 standardised hygiene indicators with k-means clusters (95% normal ellipses) and loading vectors for the seven indicators with the largest loadings. (B) Percentage of samples positive for any target pathogen (solid) and microbiologically unsatisfactory (hatched) by hygiene cluster, with Wilson 95% confidence intervals. (C) Total plate count of food samples against holding time by holding condition, with LOWESS curves.

## 4. Discussion

This study links vendor-level hygiene observations to laboratory results for street foods in a secondary town of north-eastern Bangladesh. About one in three samples carried *E. coli*, *Salmonella* spp. or *S. aureus*, four in ten were microbiologically unsatisfactory, and more than half of the isolates were multidrug resistant. Two everyday practices, failure to cover food and irregular hand washing, were independently associated with contamination after adjustment for sample type, water source and other behaviours.

The overall pathogen positivity of 35.6% falls within the range reported in other Bangladeshi studies, which have found *E. coli* and *Staphylococcus* spp. in a substantial share of fast foods and beverages in Mymensingh and Gazipur [20], and is consistent with surveys from other low- and middle-income cities [21]. The finding that water used to prepare juice was the most frequently contaminated sample type, with *E. coli* in 42.9%, is important because this water is added to a product that is not heated further. Treating or replacing the water used for beverages is therefore a direct control point. *S. aureus* in burgers and grilled chicken and coliforms on almost half of vendors’ hands point to post-cooking contamination through handling, which is the main way *S. aureus* reaches food.

Food covering had the strongest association with contamination. Uncovered food is exposed to dust, flies and the hands of customers and passers-by, and the stalls in this study were often next to open or poorly maintained drains [22]. The gradient from always to sometimes to never covering (AOR 1, 2.67 and 5.16) supports a dose–response relationship rather than a chance finding. Irregular hand washing doubled the odds of contamination, in line with the long-standing view that food handlers are a principal route of faecal and skin bacteria into ready-to-eat food [23]. Both practices are cheap to change, easy for an inspector to check, and part of the WHO Five Keys to Safer Food. Only 42.7% of stalls had a hand-washing station and 35.5% had soap, so promotion of hand washing must come with the means to do it.

Three associations ran against expectation and need careful interpretation. Samples from vendors using tube-well water were less often contaminated than those from vendors using municipal supply water. Piped supply in Bangladeshi towns is often intermittent, and low-pressure periods allow contaminated groundwater to enter leaking pipes; vendors also store supply water in open containers for long periods. Source water was not tested, so these explanations cannot be separated. Occasional glove users had lower odds of contamination than constant glove users. Gloves worn throughout the day while handling money, raw ingredients and waste may carry more bacteria than washed bare hands, and gloves can give vendors a false sense of protection. Finally, untrained vendors had lower odds than trained vendors. Training may have been short, unstructured or not followed up, and vendors with more customers or previous inspection problems may have been the ones selected for training (reverse causation). The cluster analysis supports the view that training and inspection, as currently delivered, have not translated into better microbiological outcomes: the trained and inspected cluster was no less contaminated than the untrained cluster. Knowledge was also unrelated to contamination, which matches reports from Dhaka that knowledge alone does not change practice [24,25]. These inverse findings should be treated as hypotheses for confirmation, not as evidence that gloves or training are harmful.

The AMR profile was dominated by resistance to the older, cheaper drugs used in both human and veterinary practice. Penicillin resistance was present in 81% of isolates and tetracycline resistance in more than half, consistent with previous Bangladeshi reports of high penicillin and tetracycline resistance in isolates from street foods [26,27]. The 55.6% MDR rate and the 80.8% of isolates with a MAR index above 0.2 suggest that the bacteria originate from environments with heavy antimicrobial exposure [28]. Resistance to fluoroquinolones in 26–38% of isolates, including enrofloxacin, which is used only in animals, is a concern because fluoroquinolones are first-line drugs for invasive salmonellosis and are classified by WHO as critically important. Co-resistance clustered within classes, as expected from shared mechanisms. The UpSet plot added information that drug-by-drug percentages hide: a single combination, penicillin–fluoroquinolone–tetracycline, accounted for one in seven isolates, most of them *S. aureus*, which suggests a common source or a circulating resistant lineage. Molecular typing and detection of resistance genes would be needed to test this.

The spatial analysis did not show significant differences in pathogen positivity between sites, but mean TPC did differ, and the MDR surface identified the Bus Stand area as a possible hotspot. Travellers who eat at the bus stand disperse quickly, so illness acquired there is unlikely to be detected locally. These site estimates rest on few isolates and should guide targeted sampling rather than conclusions.

The study has limitations. It is cross-sectional and covers three post-monsoon months, so seasonal variation and causality cannot be assessed. Sampling sites were mapped with approximate centroids, and the number of sites was too small for formal spatial statistics. The multivariable model had about 5.6 events per parameter, below the conventional ten, although simulation work suggests that five to nine events per variable usually give acceptable estimates [29,30]; the wide confidence intervals for some categories reflect this. Practices were observed on single visits and may have changed in the observers’ presence. Isolate confirmation was phenotypic, and resistance genes, virulence factors and enterotoxin production were not examined.

## 5. Conclusions

Street-vended foods and the water used to prepare them in Habiganj are frequently contaminated with *E. coli*, *S. aureus* and *Salmonella* spp., and more than half of these isolates are multidrug resistant. Keeping food covered and washing hands regularly were the modifiable practices most strongly associated with lower contamination. Municipal programmes should provide covered display units, hand-washing stations with soap and safe water for beverages, and should evaluate whether current training and inspection actually change practice. Street foods in secondary towns should be included in Bangladesh’s integrated AMR surveillance.

## Declarations

### Author contributions

Sharmin Sultana Royal (Conceptualization, Methodology, Investigation, Project administration, Writing – original draft). Salauddin Yousup (Investigation, Data curation). Md. Abul Kalam Azad (Investigation, Data curation). Md. Shamim Uddin (Investigation). Tasnim Zarin (Investigation, Validation). FNU Nahiduzzaman (Formal analysis, Visualization, Writing – original draft). Md. Ali Asgar (Methodology, Writing – review & editing). Md. Ariful Islam (Conceptualization, Project Admisinitration, Funding, Supervision, Writing – review & editing). Md. Azizul Haque (Conceptualization, Supervision, Writing – review & editing).

### Ethics and informed consent

This study was conducted as a collaborative research initiative involving Bangladesh Agricultural University (BAU), Mymensingh, and Habiganj Agricultural University (HAU), Habiganj, Bangladesh, under the umbrella of a broader research project entitled “Integrated Assessment of Bacterial Contamination, Transmission Pathways, and Predictive Risk in Street-Vended Foods in Bangladesh.” The overarching project received ethical approval from the Animal Welfare and Experimentation Ethics Committee (AWEEC) of Bangladesh Agricultural University (Approval No. AWEEC/BAU/2024(2)/30(a); dated 12 December 2024). The Habiganj-specific study protocol was also documented by the Research Ethics Committee of Habiganj Agricultural University (Approval No. HAU-REC-EAC-000012; dated 4 May 2026). All research activities were conducted in accordance with applicable institutional ethical guidelines. Informed consent was obtained from all participating vendors before interviews and sample collection. Participant anonymity and confidentiality were maintained throughout the study, and all collected information was used exclusively for scientific research purposes.

### Competing interests

The authors declare no competing interests.

### Data availability

The de-identified dataset and the analysis code are available from the corresponding author on reasonable request.

## Acknowledgements

The authors thank the street-food vendors of Habiganj municipality who took part in the study.

## Notes

### Competing Interest Statement

The authors have declared no competing interest.

### Author Declarations

The study was conducted as a collaborative research initiative involving Bangladesh Agricultural University (BAU), Mymensingh, and Habiganj Agricultural University (HAU), Habiganj, Bangladesh, under the umbrella of the broader research project entitled “Integrated Assessment of Bacterial Contamination, Transmission Pathways, and Predictive Risk in Street-Vended Foods in Bangladesh.” The overarching project received ethical approval from the Animal Welfare and Experimentation Ethics Committee (AWEEC), Bangladesh Agricultural University (Approval No. AWEEC/BAU/2024(2)/30(a) dated 12 December 2024). The Habiganj-specific study protocol was additionally documented by the Research Ethics Committee of Habiganj Agricultural University (Ref. No. HAU/HAUREC/EAL-11/2026-27/2026 dated 4 May 2026). Informed consent was obtained from all participating vendors before interviews and sample collection, and participant confidentiality was maintained throughout the study.

